# Building an Auditable Reference Standard for Clinical Information Extraction from Transesophageal Echocardiography Reports

**DOI:** 10.64898/2026.11.19.26368500

**Authors:** Emily J. MacKay, Michael S. Kaplan, Mingyang Yuan, Annie L. Chen, Yujia Ge, Layla Canales, Emily Yu, Rohan Shad, Bo Zhang, Alison M. Pouch

**Affiliations:** Department of Anesthesiology and Critical Care, Perelman School of Medicine at the University of Pennsylvania, Philadelphia, PA, United States; Department of Anesthesiology and Pain Medicine, University of Washington, Seattle, WA, United States; Lewis Katz School of Medicine, Temple University, Philadelphia, PA, United States; Perelman School of Medicine, University of Pennsylvania, Philadelphia, PA, United States; Tufts University School of Medicine, Boston, MA, United States; Villanova University, Villanova, PA, United States; State University of New York Downstate Health Sciences University, New York City, NY, United States; Division of Cardiovascular Surgery, Perelman School of Medicine at the University of Pennsylvania, Philadelphia, PA, United States; Vaccine and Infectious Disease Division, Fred Hutchinson Cancer Center, Seattle, WA, United States; Department of Radiology, University of Pennsylvania, Philadelphia, PA, United States; Department of Bioengineering, University of Pennsylvania, Philadelphia, PA United States

## Abstract

**Objectives:** To develop an auditable, two-institution reference standard for intraoperative transesophageal echocardiography (TEE) reports and evaluate whether reviewer disagreement identifies limitations in the underlying clinical information model.

**Materials and Methods:** We selected 300 reports (150 per institution) from 925 at the University of Pennsylvania and University of Washington using random sampling and enrichment for challenging documentation. Four trained abstractors applied a shared 39-variable V1 codebook. Independent paired comparisons included 297 reports; three historical reports contributed only to the final reference standard. Categorical agreement used exact agreement and unweighted Cohen’s *κ*; numeric left-ventricular ejection fraction (LVEF) used exact agreement and mean absolute differences. Disagreements underwent structured expert adjudication and reason classification. Additional analyses assessed reviewer-pair, sampling-stratified, and population-weighted agreement.

**Results:** Among 11,583 formal reviewer-comparison cells, 10,293 (88.9%) were resolved by reviewer agreement and 1,290 (11.1%) required targeted expert adjudication (1,289 tasks). Codebook/schema ambiguity (40.8%) and interpretation/classification differences (25.2%) together accounted for 66.0% of targeted tasks. Adjudication was concentrated in aortic dissection, valve type, and post-intervention findings. Categorical *κ* ranged from 0.364 to 0.960 and was inversely correlated with targeted-adjudication burden (Spearman ρ=−0.884; 95% CI, −0.955 to −0.734). Sampling-weighted *κ* changed by no more than 0.05 for 32 of 35 categorical variables.

**Discussion:** Structured adjudication identified recurring schema and interpretation challenges beyond individual abstraction errors, providing specific targets for information-model refinement.

**Conclusion:** As NLP and LLM-based abstraction tools move toward clinical use, the reference standards used to evaluate them require comparably rigorous development, adjudication, and independent validation.

## Introduction

Clinical information extraction is becoming increasingly reliant on natural language processing and large language models (LLMs) to transform free-text found in narrative reports into structured data.[1 2] However, model performance is only as meaningful as the reference standard used to define a correct extraction.[3] The recently published Clinical Information Extraction Reporting Guideline (CINEX) emphasizes that rigorous evaluation requires an explicit clinical information model, transparent annotation rules, assessment of inter-annotator agreement, and systematic handling of disagreement.[2 4] These requirements become particularly important when the target is not a simple entity or measurement, but a clinically interpreted parameter whose meaning depends on context, temporality, missingness, and relationships among findings.[5 6] In such settings, reviewer disagreement may reflect more than annotation error: it may expose ambiguity or under specification in the information model itself.[7 8] Yet annotation guidelines are rarely published, and disagreement is commonly resolved through adjudication without being analyzed as a source of methodological information.[9]

Intraoperative transesophageal echocardiography (TEE) provides a demanding test case for this problem. A single report may describe multiple cardiac structures before and after cardiopulmonary bypass or surgical intervention; combine quantitative measurements with qualitative assessments; document transient physiologic abnormalities that subsequently resolve; describe prosthetic or surgically altered anatomy; and distinguish findings that are absent, unchanged, unable to be assessed, or simply not mentioned.[10 11] The same clinical concept may also be represented differently across institutions.[12] These features create dependencies that are difficult to represent with simple extraction rules. A post-intervention finding may or may not legitimately inherit a pre-intervention state; a numeric ejection fraction may coexist with a distinct qualitative assessment; and the interpretation of “normal” may depend on whether it refers to structure, function, or the absence of clinically significant disease.[11 13] Although prior work has demonstrated that echocardiographic findings can be extracted from free-text reports using NLP and LLM-based approaches,[12–14] extending these methods from a limited set of predefined variables to comprehensive report-level information extraction requires a substantially more explicit and auditable representation of the underlying clinical concepts.

Our group previously demonstrated that LLM-based methods could extract selected perioperative TEE findings from narrative reports using a manually constructed reference standard.[15] However, comprehensive abstraction of an entire intraoperative TEE report requires substantially greater precision in how clinical concepts, temporal relationships, missingness, and inferential rules are defined. Rather than treating an initial codebook as fixed, we viewed reference-standard development as an empirical process in which disagreement could reveal where the information model was insufficiently specified. We therefore constructed a two-institution reference standard for structured clinical information extraction from intraoperative TEE reports and systematically analyzed reviewer disagreement and expert adjudication to quantify reliability, identify recurrent sources of disagreement, and determine which concepts generated the greatest expert-review burden. Our aim was not only to establish a V1 reference standard, but to use its observed failures to define the requirements for a more rigorous and prospectively specified next-generation codebook before downstream automated abstraction.[2 8]

## Methods

### Study Design and Report Sample

We conducted a two-institution, reference-standard development study using intraoperative transesophageal echocardiography (TEE) reports from the University of Pennsylvania (UPenn) and the University of Washington (UW). The source dataset comprised 925 reports (500 UPenn and 425 UW). We selected 300 reports for abstraction, balanced equally by institution. At each site, we first identified 50 reports with the highest scores on a rule-based screening approach designed to identify clinically or structurally challenging documentation patterns (see **Code and Data Availability** for repository information). An additional 100 reports were then randomly selected from the remaining reports at each institution. This deliberate enrichment was intended to stress-test the abstraction framework. Accordingly, adjudication burden was considered a characteristic of the constructed reference-standard sample rather than an estimate of disagreement frequency in the underlying clinical population. Three historical example reports predated the formal reviewer-comparison workflow. These reports were retained in the final reference standard but excluded from formal inter-rater reliability analyses, leaving 297 reports for paired reviewer comparison. The study was determined to be exempt from institutional review board review and was conducted under a fully executed data use agreement between UW and UPenn.

### Reference-standard Variables and Abstraction

Building on our prior TEE extraction work, we developed an expanded V1 codebook incorporating additional and more complex clinical concepts.[15] The present study focused on 39 variables selected for formal dual abstraction and expert adjudication. These included left-and right-ventricular size and systolic function, numeric left-ventricular ejection fraction (LVEF), regional wall-motion abnormalities, valvular structure and function, aortic dissection, pericardial effusion, and report-level timepoint availability. Most clinical variables were represented separately before and after surgical intervention.

Four abstractors (three medical students and one undergraduate student) were trained using the same V1 codebook, definitions, and structured abstraction workflow. One medical student served as the primary abstractor and reviewed all 300 reports. The three secondary abstractors each independently reviewed 100 nonoverlapping reports. Secondary reviewer assignments were nonrandom and reflected anticipated report complexity and reviewer experience. Each of the 297 reports included in formal reviewer comparison therefore had two independent pre-adjudication abstractions.

### Expert Adjudication

Reviewer disagreements were identified programmatically following independent abstraction. Formal reviewer comparison generated 1,284 disagreement tasks; five additional tasks were identified during data cleaning and quality control, yielding 1,289 targeted expert-adjudication tasks encompassing 1,290 report-variable cells. All targeted tasks were adjudicated by an expert cardiothoracic anesthesiologist with more than eight years of clinical experience in intraoperative TEE (EJM), who reviewed the source report, both reviewer responses, and applicable codebook rules before assigning the final value. The three historical example reports were reviewed separately at the full-report level.

For each targeted adjudication task, the primary reason for adjudication was classified as: (a) codebook/schema ambiguity; (b) interpretation/classification difference; (c) missed finding or omission; (d) ambiguous or insufficient report documentation; (e) data-entry or typographical error, or (f) other. “Codebook/schema” ambiguity was assigned when adjudication identified a limitation in the V1 codebook, including insufficiently specified definitions, classification options, or abstraction rules. Whereas “Interpretation/classification” differences were assigned when the existing codebook provided sufficient guidance to resolve the disagreement, but reviewers differed in applying those rules to the report. When computable, the adjudicated value was also classified as matching the primary reviewer, matching the secondary reviewer, or matching neither reviewer.

### Reference-Standard Assembly and Quality Control

The final reference standard contained 11,700 report-variable cells (300 reports × 39 variables). Each cell retained its final value and provenance, classified as reviewer agreement, targeted expert adjudication, or legacy full-report expert review. This cell-level provenance provided an auditable record of how each final value was established, allowing reviewer agreement to be distinguished from values requiring expert resolution. Automated quality-control checks confirmed the expected report, variable, site, and provenance counts and verified that no unresolved cells remained. The finalized reference standard and cell-level provenance files were frozen before analysis, establishing a locked V1 dataset for subsequent analyses.

### Statistical Analysis

Reference-standard construction and targeted expert-adjudication burden were summarized using descriptive statistics overall and by variable, clinical module, timepoint, and institution. For targeted-adjudication percentages, the denominator was the set of formal reviewer-comparison cells, excluding cells from the three legacy full-report abstractions by EJM. Adjudication reasons and reviewer-match outcomes were analyzed at the task level to avoid double-counting the single multi-variable LVEF adjudication. Site- and module-level comparisons were descriptive; formal between-site hypothesis testing was not undertaken because these comparisons were exploratory and were not a primary objective of reference-standard development.

Pre-adjudication agreement was evaluated across pooled primary-versus-assigned-secondary reviewer comparisons. For the 35 categorical variables, agreement was summarized using exact agreement and unweighted Cohen’s *κ*. Pooled *κ* estimates were calculated directly from all eligible paired reviewer observations, rather than by averaging reviewer-pair-specific estimates. These estimates characterize agreement within the implemented dual-abstraction workflow, in which one primary reviewer assessed all reports and the assigned secondary reviewer varied across reports. Exact-agreement proportions were reported with 95% Wilson score confidence intervals.[16] For the four numeric LVEF variables, agreement was summarized using exact agreement and mean absolute differences between paired reviewer values, expressed in LVEF percentage points. Agreement calculations included only reports with values recorded by both reviewers. Ninety-five percent confidence intervals for *κ* were obtained using report-level nonparametric percentile bootstrap resampling with 5,000 resamples and a fixed random seed.[17]

To characterize variation across reviewer pairs, unweighted Cohen’s *κ* was calculated separately for each primary–secondary reviewer pair, with 95% confidence intervals obtained using 5,000 report-level bootstrap resamples. To estimate categorical agreement in the 925-report source population, we recalculated pooled *κ* using inverse-probability-of-selection weights (1.0 for certainty-selected challenging reports, 4.50 for randomly sampled UPenn reports, and 3.75 for randomly sampled UW reports). Weighted *κ* was calculated using weighted observed agreement and category marginals, and estimates were compared descriptively with the primary pooled, unweighted estimates. To examine variation by institution and report complexity, unweighted Cohen’s *κ* was calculated separately for complex and randomly selected reports within each institution, with 95% confidence intervals obtained using 5,000 paired-report bootstrap resamples.

Among the 35 categorical variables, the association between pre-adjudication Cohen’s *κ* and subsequent targeted-adjudication burden was assessed using Spearman rank correlation. Numeric LVEF variables were excluded because *κ* was not calculated for these variables. The 95% confidence interval for the Spearman coefficient was estimated using nonparametric percentile bootstrap resampling of the 35 variable-level pairs with 5,000 resamples and a fixed random seed.[17] Analyses were performed in Python using pandas, SciPy, and Matplotlib. Reporting was informed by applicable elements of the Clinical Information Extraction Reporting Guideline (CINEX)[2] and the Guidelines for Reporting Reliability and Agreement Studies (GRRAS).[18]

## Results

### Reference-Standard Construction

The final reference standard included 300 reports and 39 variables, yielding 11,700 report-variable cells. Three historical example reports were excluded from formal reviewer comparison, leaving 11,583 cells across 297 reports. Of these, 10,293 cells (88.9%) were resolved by reviewer agreement, while 1,290 (11.1%) required targeted expert adjudication. At least one cell required targeted adjudication in 283 of 297 reports (95.3%), with a median of 4 targeted-adjudicated cells per report (IQR, 2–6). The 1,290 cells corresponded to 1,289 adjudication tasks because one task jointly resolved the lower and upper bounds of an LVEF range. An additional 117 cells from the three historical reports were resolved by full-report expert review. No unresolved cells remained in the final reference standard (**Table 1**).

**Table 1.** V1 Reference Standard Construction and Adjudication Summary.

| Section | Characteristic | n | Denominator | % |
|---|---|---|---|---|
| Reference-standard sample | Reports | 300 |  |  |
| Reference-standard sample | UPenn reports | 150 | 300 | 50.0 |
| Reference-standard sample | UW reports | 150 | 300 | 50.0 |
| Reference-standard sample | Variables per report | 39 |  |  |
| Reference-standard sample | Frozen report-variable cells | 11700 |  |  |
| Formal reviewer comparison | Reports included in formal reviewer comparison | 297 |  |  |
| Formal reviewer comparison | Formal reviewer-comparison cells | 11583 |  |  |
| Formal reviewer comparison | Cells resolved by reviewer agreement | 10293 | 11583 | 88.9 |
| Formal reviewer comparison | Cells requiring targeted expert adjudication | 1290 | 11583 | 11.1 |
| Formal reviewer comparison | Targeted expert-adjudication tasks | 1289 |  |  |
| Expert review and final reference standard | Reports requiring targeted expert adjudication | 283 | 297 | 95.3 |
| Expert review and final reference standard | Targeted-adjudicated cells per formal report, median (IQR) | 4 (2–6) |  |  |
| Expert review and final reference standard | Legacy reports reviewed in full | 3 |  |  |
| Expert review and final reference standard | Legacy full-report expert-reviewed cells | 117 | 11700 | 1.0 |
| Expert review and final reference standard | Reports receiving any expert review | 286 | 300 | 95.3 |
| Expert review and final reference standard | All expert-resolved cells | 1407 | 11700 | 12.0 |
| Expert review and final reference standard | Unresolved cells in frozen reference standard | 0 | 11700 | 0.0 |

**Note.** Formal reviewer-comparison percentages use the 11,583 report-variable cells from the 297 reports included in formal IRR. Reviewer agreement and targeted expert adjudication therefore partition the same formal comparison denominator (88.9% and 11.1%, respectively). The three legacy reports were reviewed in full and are reported separately. The median and IQR of targeted-adjudicated cells are calculated across all 297 formal-review reports, including reports with zero targeted adjudications. One targeted multi-variable LVEF task resolved two report-variable cells, so the number of targeted adjudication tasks (1,289) differs from the number of targeted adjudicated cells (1,290). Percentages are descriptive of this deliberately constructed reference-standard sample and should not be interpreted as population estimates of disagreement frequency. No 95% confidence intervals are reported in Table 1 because these rows are exact dataset accounting and report-variable cells are not independent.

### Adjudication Burden

Targeted adjudication was unevenly distributed across clinical variables and modules, with the greatest burden involving aortic dissection, valve type, and post-intervention findings (**Figure 1**; **Supplemental Table S1**). The highest rates were observed for aortic dissection (29.3% post-intervention; 27.6% pre-intervention), tricuspid valve type (26.6% post-intervention; 23.2% pre-intervention), and mitral valve type (22.6% pre-intervention; 20.2% post-intervention). Other frequently adjudicated variables included post-intervention aortic valve morphology (17.8%) and left-ventricular size (17.2%). At the module level, adjudication burden was greatest for the aorta (28.5%) and tricuspid valve (16.1%; **Supplemental Table S2**). Across the 19 pre-intervention and 19 post-intervention variables, post-intervention cells required targeted adjudication more frequently than pre-intervention cells (14.7% vs 8.0%).

**Figure 1.**
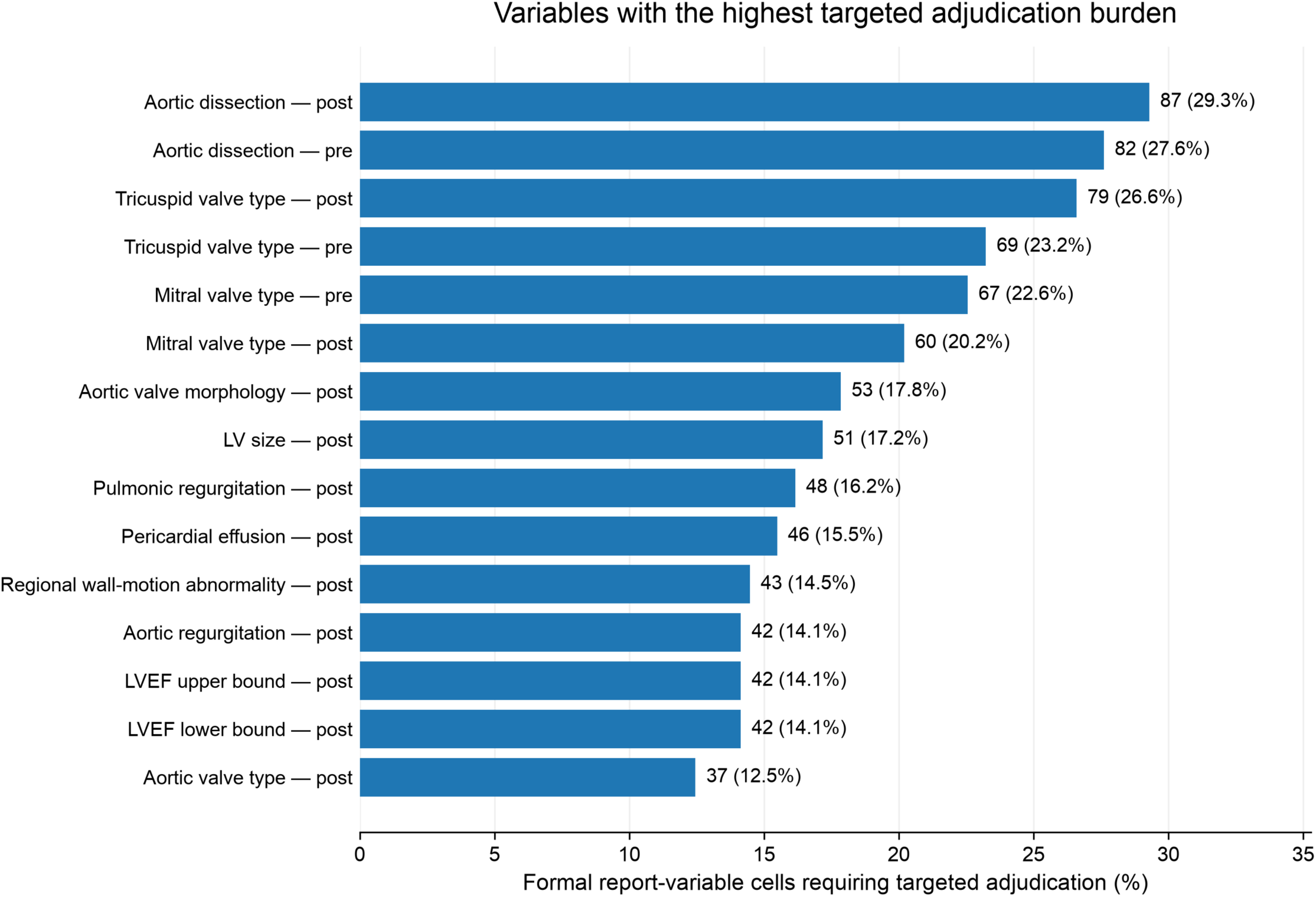
The 15 variables with the highest percentages of formal report-variable cells requiring targeted expert adjudication. Bars indicate the percentage of cells requiring adjudication, with corresponding counts and percentages displayed. Pre- and post-intervention variables are presented separately. Percentages use the 297 reports included in formal reviewer comparison as the denominator for each variable.

### Disagreement Reasons

Among 1,289 targeted adjudication tasks, the most common reasons were codebook/schema ambiguity (40.8%) and interpretation/classification differences (25.2%), which together accounted for 66.0% of targeted adjudications (**Figure 2**; **Supplemental Table S3**). Other reasons included missed findings or omissions (16.9%), ambiguous or insufficient report documentation (8.7%), data-entry or typographical errors (6.4%), and other causes (1.9%). Among 1,251 tasks for which reviewer matching was computable, the expert-adjudicated value matched the secondary reviewer in 633 (50.6%), the primary reviewer in 548 (43.8%), and neither reviewer in 70 (5.6%).

**Figure 2.**
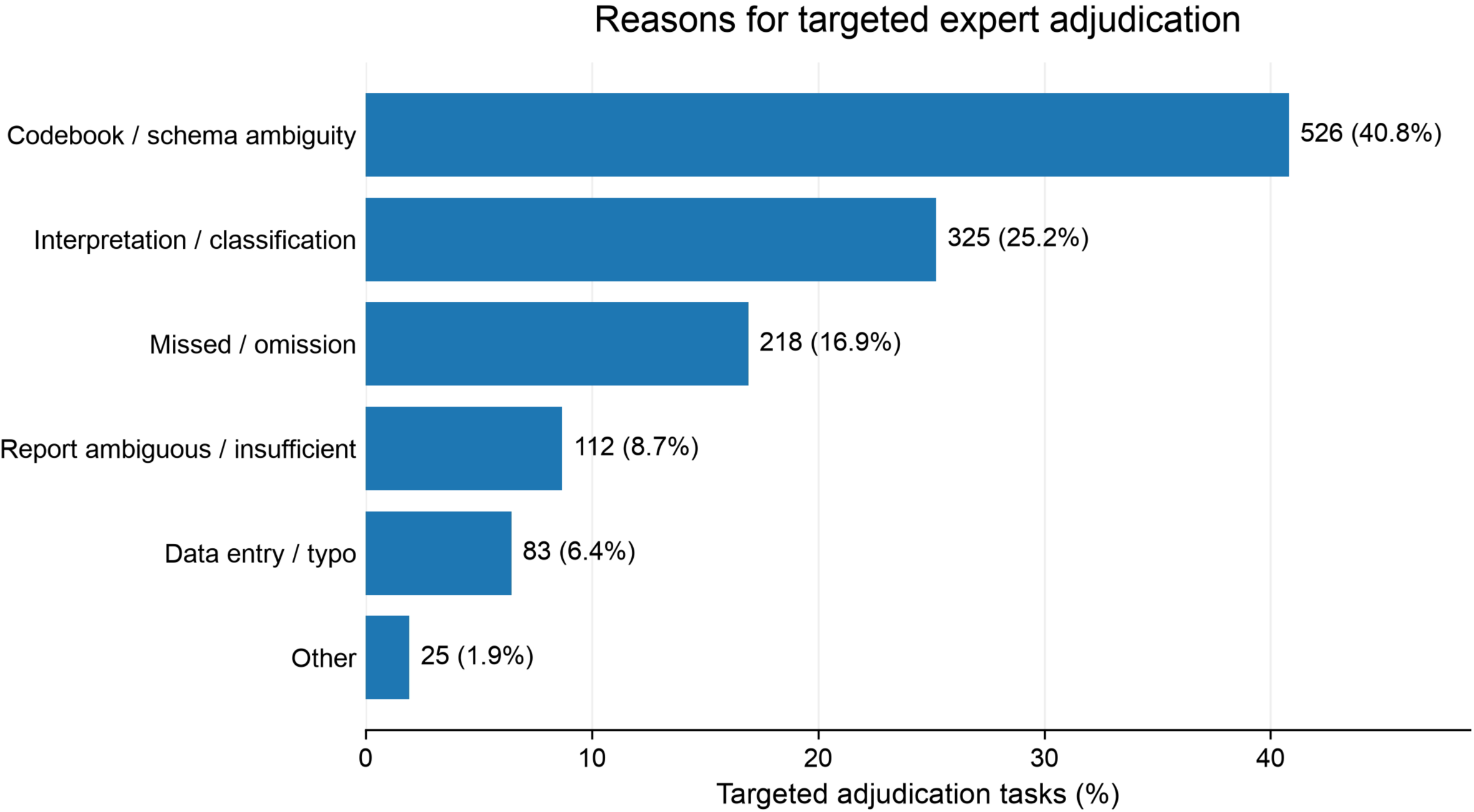
Reasons for targeted expert adjudication. Distribution of the primary reason assigned to each of 1,289 targeted expert-adjudication tasks. Each task was classified into one mutually exclusive category. Percentages use all 1,289 targeted tasks as the denominator.

### Reliability

Pre-adjudication agreement varied substantially across categorical variables (**Supplemental Table S1**). Cohen’s *κ* ranged from 0.364 to 0.960, with the lowest estimates observed for pre-intervention mitral-valve type (*κ*=0.364; 95% CI, 0.257– 0.471), post-intervention aortic dissection (*κ*=0.405; 95% CI, 0.310–0.500), pre-intervention aortic dissection (*κ*=0.454; 95% CI, 0.363–0.549), and post- and pre-intervention tricuspid-valve type (*κ*=0.529 and 0.558, respectively). For the four numeric LVEF variables, exact agreement among paired numeric values ranged from 98.7% to 100%, with mean absolute differences of 0.00–0.12 LVEF percentage points.

Reviewer-pair-specific *κ* estimates demonstrated variability for selected categorical variables, particularly valve-type classifications (**Supplemental Table S4**). Because secondary reviewer assignments were nonrandom and informed by anticipated case complexity and reviewer experience, these comparisons are considered descriptive rather than measures of individual reviewer performance. With inverse-probability weighting, *κ* changed by no more than 0.05 for 32 of 35 categorical variables (91.4%), with a median absolute change of 0.013. The largest decreases were observed for tricuspid-valve type and pre-intervention aortic dissection (**Supplemental Table S5**). Across the 35 categorical variables, lower Cohen’s *κ* was inversely correlated with targeted-adjudication burden (Spearman *ρ*=−0.884; 95% CI, −0.955 to −0.734; **Figure 3**).

**Figure 3.**
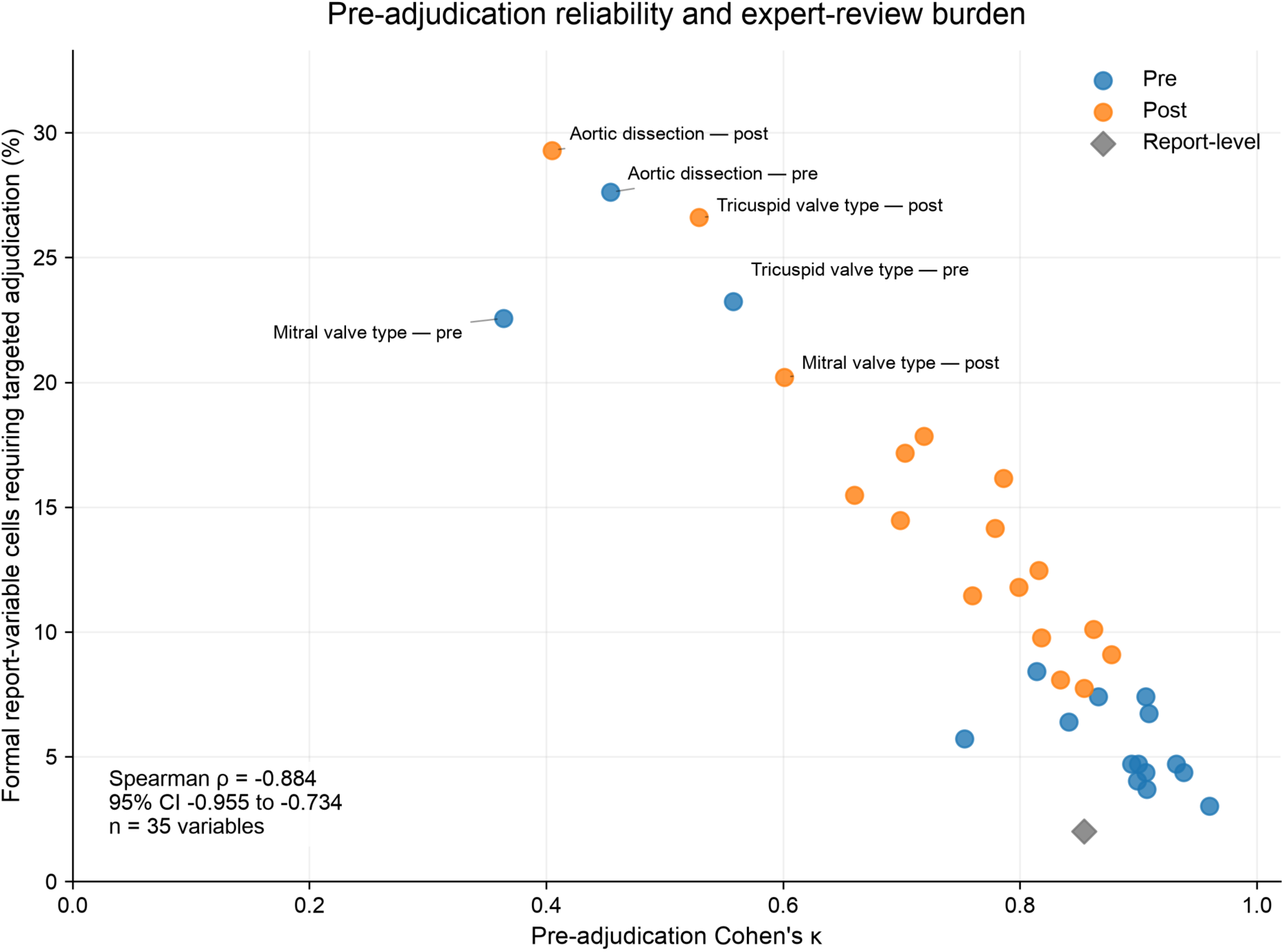
Each point represents one of 35 categorical variables, plotted by Cohen’s κ and the percentage of formal report-variable cells requiring targeted expert adjudication. Colors distinguish pre- and post-intervention variables. Spearman rank correlation and its 95% confidence interval are shown.

### Adjudication Patterns Across Institutions

Overall targeted-adjudication burden was 11.3% at UPenn and 11.0% at UW, with variation across clinical categories (**Figure 4**; **Supplemental Table S2**). Aortic variables required targeted adjudication in 16.7% of UPenn and 40.0% of UW formal report-variable cells, whereas tricuspid-valve variables required adjudication in 22.3% and 10.0%, respectively. For tricuspid-valve type, codebook/schema ambiguity was the predominant reason for adjudication at both institutions (91.8% at UPenn and 94.7% at UW).

**Figure 4.**
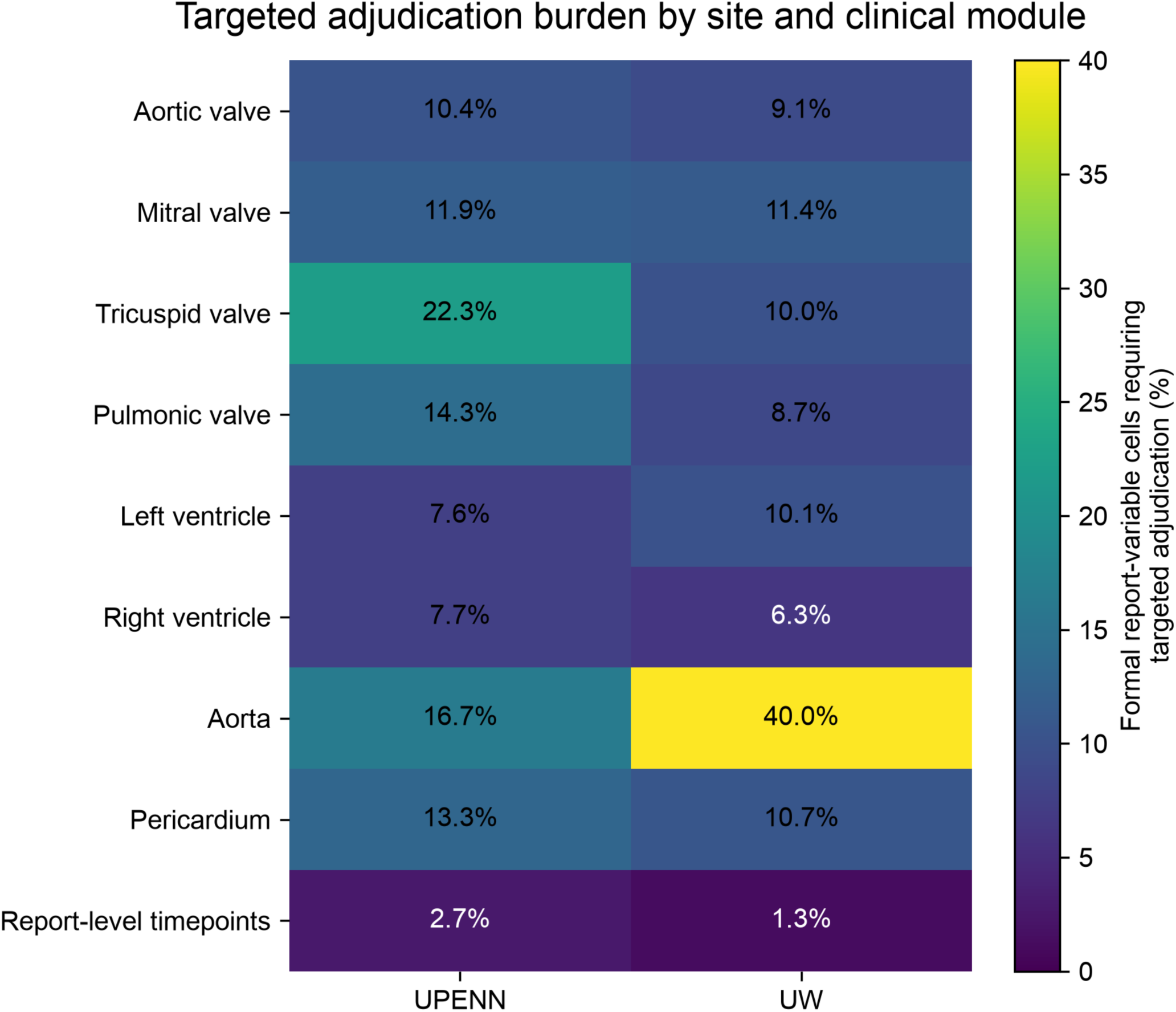
Heatmap showing the percentage of formal report-variable cells requiring targeted expert adjudication across clinical modules at the University of Pennsylvania (UPenn) and University of Washington (UW). Cell values represent the proportion of adjudicated cells within each institution and clinical module, with darker-to-lighter colors indicating increasing adjudication burden. Percentages are descriptive and use institution- and module-specific formal reviewer-comparison cells as denominators.

In analyses stratified by institution and sampling group, categorical reliability estimates varied across clinical variables, with no consistent pattern according to report complexity (**Supplemental Table S6**). For pre-intervention aortic dissection, *κ* estimates were 0.480 (UPenn complex), 0.659 (UPenn random), 0.840 (UW complex), and 0.112 (UW random). These findings are descriptive and may reflect variation in case mix, category prevalence, and nonrandom reviewer assignment.

## Discussion

In this two-institution study, development of an auditable reference standard revealed important limitations in the clinical information model for intraoperative TEE. Although most report-variable cells were resolved by reviewer agreement, targeted adjudication was concentrated in particular clinical concepts, including aortic dissection and valve type, and was more frequent for post-intervention findings. Two-thirds of targeted adjudications reflected codebook/schema ambiguity or differences in interpretation and classification, rather than simple omission or data-entry error. Variables with lower pre-adjudication reliability also required more expert review. These challenges are particularly important in intraoperative TEE, where a single report integrates serial assessments, procedural interventions, altered anatomy, and transient findings rather than describing a single time-point examination. Together, these findings suggest that systematic analysis of reviewer disagreement can identify limitations in the underlying information model, in addition to resolving individual report-variable cells.[8]

Disagreement in clinical abstraction is often treated primarily as a reviewer-performance problem, although inconsistent labels may also reflect unclear guidelines, insufficient source information, or ambiguity in how a clinical concept should be represented.[3] Our findings support this broader interpretation. While some disagreements reflected missed findings or data-entry errors, most arose from codebook/schema ambiguity or differences in mapping report language to predefined categories. For example, disagreement may arise when a pre-intervention finding is not explicitly reassessed after surgery, leaving reviewers to determine whether the earlier classification should be carried forward or considered not documented. Related challenges have been described in pathology-report extraction, where 39.4% of inter-annotator disagreements were attributed to inherent ambiguity in source reports rather than human error,[7] and in error ontologies that distinguish schema limitations from annotation and model errors.[8] The inverse association between pre-adjudication Cohen’s *κ* and expert-review burden further identified variables warranting closer scrutiny during information-model development, although both measures reflect underlying reviewer disagreement. Adjudication therefore served two functions: resolving individual report-variable cells and identifying potential systematic weaknesses in the underlying representation.

Evaluation across institutions is also important during information-model development. Heterogeneous documentation practices are a well-recognized barrier to NLP portability across institutions,[19] including in transthoracic echocardiography.[12] Although overall targeted-adjudication burden was similar at UPenn and UW, its distribution across clinical modules differed. Aortic variables required more frequent adjudication at UW, whereas tricuspid-valve type generated substantial schema-related disagreement at both institutions. These observations suggest that evaluating a shared information model across different reporting environments may reveal limitations that are not apparent within a single institution. Aggregate measures of agreement or adjudication burden may obscure these domain-specific patterns.

These findings have important implications for automated clinical information extraction. Recent NLP and LLM-based approaches have demonstrated promising performance in extracting structured findings from narrative clinical reports, including transthoracic[12 20] and transesophageal[15] echocardiography reports. However, measured extraction performance is necessarily conditional on the reference standard used to define correctness.[21] An extraction system may reproduce a label accurately even when the underlying clinical concept is incompletely specified.[22] This concern is particularly relevant to intraoperative TEE, where interpretation depends on temporality, procedural context, missingness, altered anatomy, and surgical interventions.[10 23] In such settings, rigorous and auditable reference-standard development, with preserved provenance for individual abstraction decisions and transparent expert adjudication, is an essential upstream requirement for evaluating automated extraction systems.[2 24]

The V1 adjudication process directly informed refinement of the abstraction framework, particularly the handling of temporality, missingness, inheritance of prior findings, and complex structural findings. The goal of these refinements is to reduce reliance on case-by-case expert interpretation by specifying clinical concepts and abstraction rules more precisely, while preserving expert adjudication for genuinely ambiguous source documentation. However, evaluating the revised framework using the same reports would not provide an independent assessment of its reliability. The next step, therefore, will be to specify and freeze the V2 information model before applying it to an independent set of reports, using independent abstraction, predefined agreement measures, structured adjudication, and preserved cell-level provenance.[2] Separating information-model development from independent validation is particularly important before the resulting reference standard is used to evaluate downstream NLP or LLM extraction systems.[25]

These findings should be interpreted in the context of several methodological limitations. First, the study included only two institutions and focused on the specialized domain of intraoperative TEE, potentially limiting generalizability to other clinical reporting environments. Second, the reference-standard sample was deliberately enriched for challenging reports. Stratified analyses also demonstrated variable-specific differences in reliability without a consistent relationship to report complexity; these descriptive comparisons may reflect differences in category prevalence, case mix, and reviewer assignment rather than complexity alone. Although sampling-weighted *k* estimates were generally similar to unweighted estimates, observed adjudication frequencies and disagreement-reason distributions should not be interpreted as representative of the underlying clinical population. Third, secondary abstraction was distributed across three reviewers examining nonoverlapping report subsets, with assignments informed by anticipated case complexity and reviewer experience. Consequently, reviewer-specific agreement estimates cannot be interpreted as direct comparisons of reviewer performance, and the pooled estimates reflect the observed reviewer assignment structure rather than agreement between interchangeable raters. Fourth, expert adjudication relied on the clinical judgment of a single experienced adjudicator, including classification of the primary reason for disagreement. These classifications provide a structured characterization of disagreement but do not independently establish its underlying cause. Because targeted adjudication was limited to reviewer disagreements, concordant but incorrect interpretations by both reviewers would not have been identified, and reviewer agreement should not be interpreted as independent confirmation of clinical accuracy. Three historical reports were also reviewed outside the formal reviewer-comparison workflow. Fifth, the observed association between Cohen’s *κ* and adjudication burden should be interpreted descriptively, as both measures reflect the same underlying reviewer disagreements. Finally, this study evaluated reference-standard development rather than automated extraction performance, and the refined V2 information model has not yet undergone independent validation.

## Conclusions

Reference standards for automated clinical information extraction are often treated as fixed ground truth, with limited attention to how disagreement was identified and resolved. In intraoperative TEE, systematic analysis of reviewer disagreement and expert adjudication revealed important limitations in the clinical information model and provided a structured basis for its refinement. As NLP and LLM-based abstraction tools move toward clinical use, the reference standards used to evaluate them must be developed, adjudicated, and independently validated with comparable methodological rigor.

## Code and Data Availability

A public repository for this study is available through the TEECompass GitHub organization (https://github.com/TEECompass/tee-reference-standard-v1). Analysis code is being prepared for release, subject to institutional review of licensing and intellectual property. Patient-level clinical reports and adjudication data are not publicly available due to institutional data-use agreements and privacy protections.

## Disclosures

This study was funded, in-part, by: (a) The National Institutes of Health (NIH), National Heart, Lung, and Blood Institute (NHLBI) [grant number: K23HL166964 to E.J.M.]

## Competing Interests

The authors declare no competing interests.

## Data Availability

The clinical reports and patient-level adjudication data used in this study are not publicly available due to institutional data-use agreements and patient privacy protections. The study was conducted using data from the University of Pennsylvania and the University of Washington. A public repository for associated analysis code has been established at https://github.com/TEECompass/tee-reference-standard-v1; code release is pending institutional review.

## Notes

### Competing Interest Statement

The authors have declared no competing interest.

### Author Declarations

The study was determined to be exempt from institutional review board (IRB) review by both the University of Pennsylvania (UPenn) and the University of Washington (UW), and was conducted under a fully executed data use agreement between both institutions.

