## Supplementary Materials for "Building an Auditable Reference Standard for Clinical Information Extraction from Transesophageal Echocardiography Reports"

### Table S1. Variable-Level Pre-Adjudication Reliability and Targeted Adjudication Burden

| Variable | Variable code | Module | Timepoint | Field type | IRR n | Exact agreement % (95% CI) | Pooled Cohen's κ (95% CI) | LVEF mean absolute difference (pp) | Formal cells | Targeted adjudication n | Targeted adjudication % |
|---|---|---|---|---|---|---|---|---|---|---|---|
| Aortic valve morphology | pre_av_morphology | Aortic valve | Pre | categorical | 297 | 92.6 (89.0–95.1) | 0.866 (0.811–0.914) | | 297 | 22 | 7.4 |
| Aortic regurgitation | pre_av_regurgitation | Aortic valve | Pre | categorical | 297 | 92.6 (89.0–95.1) | 0.906 (0.867–0.940) | | 297 | 22 | 7.4 |
| Aortic stenosis | pre_av_stenosis | Aortic valve | Pre | categorical | 297 | 95.3 (92.2–97.2) | 0.932 (0.897–0.965) | | 297 | 14 | 4.7 |
| Aortic valve type | pre_av_type | Aortic valve | Pre | categorical | 297 | 94.3 (91.0–96.4) | 0.753 (0.640–0.854) | | 297 | 17 | 5.7 |
| Aortic valve morphology | post_av_morphology | Aortic valve | Post | categorical | 297 | 82.2 (77.4–86.1) | 0.719 (0.654–0.780) | | 297 | 53 | 17.8 |
| Aortic regurgitation | post_av_regurgitation | Aortic valve | Post | categorical | 297 | 85.9 (81.4–89.4) | 0.779 (0.717–0.837) | | 297 | 42 | 14.1 |
| Aortic stenosis | post_av_stenosis | Aortic valve | Post | categorical | 297 | 91.9 (88.3–94.5) | 0.834 (0.769–0.893) | | 297 | 24 | 8.1 |
| Aortic valve type | post_av_type | Aortic valve | Post | categorical | 297 | 87.5 (83.3–90.8) | 0.816 (0.760–0.866) | | 297 | 37 | 12.5 |
| Mitral regurgitation | pre_mv_regurgitation | Mitral valve | Pre | categorical | 297 | 97.0 (94.3–98.4) | 0.960 (0.932–0.982) | | 297 | 9 | 3.0 |
| Mitral stenosis | pre_mv_stenosis | Mitral valve | Pre | categorical | 297 | 96.3 (93.5–97.9) | 0.907 (0.850–0.958) | | 297 | 11 | 3.7 |
| Mitral valve type | pre_mv_type | Mitral valve | Pre | categorical | 297 | 77.4 (72.4–81.8) | 0.364 (0.257–0.471) | | 297 | 67 | 22.6 |
| Mitral regurgitation | post_mv_regurgitation | Mitral valve | Post | categorical | 297 | 90.9 (87.1–93.7) | 0.877 (0.833–0.918) | | 297 | 27 | 9.1 |
| Mitral stenosis | post_mv_stenosis | Mitral valve | Post | categorical | 297 | 88.6 (84.4–91.7) | 0.760 (0.679–0.830) | | 297 | 34 | 11.4 |
| Mitral valve type | post_mv_type | Mitral valve | Post | categorical | 297 | 79.8 (74.9–84.0) | 0.601 (0.512–0.682) | | 297 | 60 | 20.2 |

| Variable | Variable code | Module | Timepoint | Field type | IRR n | Exact agreement % (95% CI) | Pooled Cohen's κ (95% CI) | LVEF mean absolute difference (pp) | Formal cells | Targeted adjudication n | Targeted adjudication % |
|---|---|---|---|---|---|---|---|---|---|---|---|
| Tricuspid regurgitation | pre_tv_regurgitation | Tricuspid valve | Pre | categorical | 297 | 95.6 (92.7–97.4) | 0.938 (0.903–0.967) | | 297 | 13 | 4.4 |
| Tricuspid valve type | pre_tv_type | Tricuspid valve | Pre | categorical | 297 | 76.8 (71.6–81.2) | 0.558 (0.475–0.637) | | 297 | 69 | 23.2 |
| Tricuspid regurgitation | post_tv_regurgitation | Tricuspid valve | Post | categorical | 297 | 89.9 (85.9–92.8) | 0.862 (0.813–0.907) | | 297 | 30 | 10.1 |
| Tricuspid valve type | post_tv_type | Tricuspid valve | Post | categorical | 297 | 73.4 (68.1–78.1) | 0.529 (0.449–0.609) | | 297 | 79 | 26.6 |
| Pulmonic regurgitation | pre_pv_regurgitation | Pulmonic valve | Pre | categorical | 297 | 93.3 (89.8–95.6) | 0.909 (0.868–0.945) | | 297 | 20 | 6.7 |
| Pulmonic regurgitation | post_pv_regurgitation | Pulmonic valve | Post | categorical | 297 | 83.8 (79.2–87.6) | 0.786 (0.728–0.841) | | 297 | 48 | 16.2 |
| LV size | pre_lv_size | Left ventricle | Pre | categorical | 296 | 95.6 (92.6–97.4) | 0.900 (0.842–0.950) | | 297 | 14 | 4.7 |
| LV systolic function | pre_lv_systolic_function | Left ventricle | Pre | categorical | 297 | 95.3 (92.2–97.2) | 0.894 (0.837–0.945) | | 297 | 14 | 4.7 |
| LVEF upper bound | pre_lvef_high | Left ventricle | Pre | numeric | 233 | 98.7 (96.3–99.6) | | 0.12 | 297 | 5 | 1.7 |
| LVEF lower bound | pre_lvef_low | Left ventricle | Pre | numeric | 233 | 99.1 (96.9–99.8) | | 0.10 | 297 | 4 | 1.3 |
| Regional wall-motion abnormality | pre_rwma_present | Left ventricle | Pre | categorical | 296 | 93.9 (90.6–96.1) | 0.841 (0.766–0.905) | | 297 | 19 | 6.4 |
| LV size | post_lv_size | Left ventricle | Post | categorical | 296 | 83.1 (78.4–86.9) | 0.703 (0.631–0.772) | | 297 | 51 | 17.2 |
| LV systolic function | post_lv_systolic_function | Left ventricle | Post | categorical | 297 | 90.2 (86.3–93.1) | 0.818 (0.754–0.877) | | 297 | 29 | 9.8 |
| LVEF upper bound | post_lvef_high | Left ventricle | Post | numeric | 183 | 100.0 (97.9–100.0) | | 0.00 | 297 | 42 | 14.1 |
| LVEF lower bound | post_lvef_low | Left ventricle | Post | numeric | 183 | 99.5 (97.0–99.9) | | 0.05 | 297 | 42 | 14.1 |

| Variable | Variable code | Module | Timepoint | Field type | IRR n | Exact agreement % (95% CI) | Pooled Cohen's $\kappa$ (95% CI) | LVEF mean absolute difference (pp) | Formal cells | Targeted adjudication n | Targeted adjudication % |
|---|---|---|---|---|---|---|---|---|---|---|---|
| Regional wall-motion abnormality | post_rwma_present | Left ventricle | Post | categorical | 296 | 85.8 (81.4–89.3) | 0.699 (0.614–0.777) | | 297 | 43 | 14.5 |
| RV size | pre_rv_size | Right ventricle | Pre | categorical | 297 | 95.6 (92.7–97.4) | 0.906 (0.854–0.951) | | 297 | 13 | 4.4 |
| RV systolic function | pre_rv_systolic_function | Right ventricle | Pre | categorical | 297 | 96.0 (93.1–97.7) | 0.899 (0.838–0.950) | | 297 | 12 | 4.0 |
| RV size | post_rv_size | Right ventricle | Post | categorical | 296 | 88.5 (84.4–91.7) | 0.799 (0.733–0.860) | | 297 | 35 | 11.8 |
| RV systolic function | post_rv_systolic_function | Right ventricle | Post | categorical | 297 | 92.3 (88.6–94.8) | 0.854 (0.798–0.908) | | 297 | 23 | 7.7 |
| Aortic dissection | pre_aortic_dissection | Aorta | Pre | categorical | 297 | 72.4 (67.0–77.2) | 0.454 (0.363–0.549) | | 297 | 82 | 27.6 |
| Aortic dissection | post_aortic_dissection | Aorta | Post | categorical | 293 | 71.7 (66.3–76.5) | 0.405 (0.310–0.500) | | 297 | 87 | 29.3 |
| Pericardial effusion | pre_pericardial_effusion | Pericardium | Pre | categorical | 297 | 91.6 (87.9–94.2) | 0.814 (0.743–0.879) | | 297 | 25 | 8.4 |
| Pericardial effusion | post_pericardial_effusion | Pericardium | Post | categorical | 297 | 84.5 (80.0–88.2) | 0.660 (0.575–0.742) | | 297 | 46 | 15.5 |
| TEE timepoints available | tee_timepoints_available | Report-level timepoints | Report-level | categorical | 296 | 98.3 (96.1–99.3) | 0.854 (0.707–0.964) | | 297 | 6 | 2.0 |

**Note.** For categorical variables, pooled unweighted Cohen's $\kappa$ is shown with a 95% paired-report percentile bootstrap CI (5,000 resamples). Pooled $\kappa$ was calculated directly from all eligible pre-adjudication comparisons between the anchor reviewer and the assigned secondary reviewer, rather than by averaging reviewer-pair-specific $\kappa$ estimates. Secondary reviewer assignments were nonrandom and informed by anticipated case complexity and reviewer experience; thus, pooled estimates describe agreement within the implemented abstraction workflow rather than reliability between interchangeable reviewers. Reviewer-pair-specific estimates appear in Table S4. For numeric LVEF fields, exact agreement and mean absolute differences between paired values are reported, with differences expressed in LVEF percentage points (pp). Exact-agreement percentages and Wilson 95% CIs use reports with both reviewer values present. Targeted adjudication percentages use all formal report-variable cells.

### Table S2. Targeted Adjudication Burden by Clinical Module and Site

| Module | Overall formal cells | Overall targeted adjudication | UPenn targeted adjudication | UW targeted adjudication | UPenn − UW (pp) | UPenn formal cells | UW formal cells |
|---|---|---|---|---|---|---|---|
| Aortic valve | 2376 | 231 (9.7%) | 122 (10.4%) | 109 (9.1%) | 1.3 | 1176 | 1200 |
| Mitral valve | 1782 | 208 (11.7%) | 105 (11.9%) | 103 (11.4%) | 0.5 | 882 | 900 |
| Tricuspid valve | 1188 | 191 (16.1%) | 131 (22.3%) | 60 (10.0%) | 12.3 | 588 | 600 |
| Pulmonic valve | 594 | 68 (11.4%) | 42 (14.3%) | 26 (8.7%) | 5.6 | 294 | 300 |
| Left ventricle | 2970 | 263 (8.9%) | 112 (7.6%) | 151 (10.1%) | -2.4 | 1470 | 1500 |
| Right ventricle | 1188 | 83 (7.0%) | 45 (7.7%) | 38 (6.3%) | 1.3 | 588 | 600 |
| Aorta | 594 | 169 (28.5%) | 49 (16.7%) | 120 (40.0%) | -23.3 | 294 | 300 |
| Pericardium | 594 | 71 (12.0%) | 39 (13.3%) | 32 (10.7%) | 2.6 | 294 | 300 |
| Report-level timepoints | 297 | 6 (2.0%) | 4 (2.7%) | 2 (1.3%) | 1.4 | 147 | 150 |

**Note.** Percentages use formal reviewer-comparison cells as the denominator and exclude cells from the three legacy full-report reviews. Percentage-point difference is UPenn minus UW. Site-specific percentages are descriptive; no formal between-site testing was undertaken because site contrasts were exploratory and were not a primary objective of reference-standard development.

### Table S3. Adjudication Task Composition, Reasons, and Outcomes

| Section | Category | n | % | Denominator |
|---|---|---|---|---|
| Adjudication queue origin | Formal IRR disagreement | 1284 | 99.4 | 1,292 adjudication tasks |
| Adjudication queue origin | Cleaning / QC issue | 5 | 0.4 | 1,292 adjudication tasks |
| Adjudication queue origin | Legacy full-report review | 3 | 0.2 | 1,292 adjudication tasks |
| Reason for targeted adjudication | Codebook / schema ambiguity | 526 | 40.8 | 1,289 targeted tasks |
| Reason for targeted adjudication | Interpretation / classification | 325 | 25.2 | 1,289 targeted tasks |
| Reason for targeted adjudication | Missed finding / omission | 218 | 16.9 | 1,289 targeted tasks |
| Reason for targeted adjudication | Report ambiguous / insufficient | 112 | 8.7 | 1,289 targeted tasks |
| Reason for targeted adjudication | Data entry / typo | 83 | 6.4 | 1,289 targeted tasks |
| Reason for targeted adjudication | Other | 25 | 1.9 | 1,289 targeted tasks |
| Adjudication outcome | Expert adjudication matched secondary reviewer | 633 | 49.1 | 1,289 targeted tasks |
| Adjudication outcome | Expert adjudication matched anchor reviewer | 548 | 42.5 | 1,289 targeted tasks |
| Adjudication outcome | Expert adjudication matched neither reviewer | 70 | 5.4 | 1,289 targeted tasks |
| Adjudication outcome | not computable missing adjudication | 37 | 2.9 | 1,289 targeted tasks |
| Adjudication outcome | Reviewer-match outcome not computable: multi-variable item | 1 | 0.1 | 1,289 targeted tasks |
| Reviewer-match outcome among computable tasks | Expert adjudication matched secondary reviewer | 633 | 50.6 | 1,251 computable tasks |
| Reviewer-match outcome among computable tasks | Expert adjudication matched anchor reviewer | 548 | 43.8 | 1,251 computable tasks |
| Reviewer-match outcome among computable tasks | Expert adjudication matched neither reviewer | 70 | 5.6 | 1,251 computable tasks |

**Note.** Queue-origin percentages use all 1,292 adjudication tasks. Reason and overall outcome percentages use all 1,289 targeted expert-adjudication tasks as the denominator. The computable reviewer-match subset excludes tasks for which anchor-versus-secondary matching could not be computed from the adjudication record. A non-computable reviewer-match outcome does not indicate an unresolved cell in the frozen reference standard.

### Table S4. Categorical Reliability by Anchor–Secondary Reviewer Pair

| Module | Variable | A–B κ (95% CI); n | A–C κ (95% CI); n | A–D κ (95% CI); n |
|---|---|---|---|---|
| Aortic valve | Pre: Aortic valve morphology | 0.824 (0.718–0.920); n=97 | 0.884 (0.795–0.961); n=100 | 0.891 (0.795–0.975); n=100 |
| Aortic valve | Pre: Aortic regurgitation | 0.897 (0.823–0.961); n=97 | 0.910 (0.839–0.973); n=100 | 0.909 (0.837–0.972); n=100 |
| Aortic valve | Pre: Aortic stenosis | 0.911 (0.835–0.971); n=97 | 0.899 (0.821–0.969); n=100 | 0.985 (0.951–1.000); n=100 |
| Aortic valve | Pre: Aortic valve type | 0.740 (0.547–0.888); n=97 | 0.877 (0.731–1.000); n=100 | 0.522 (0.117–0.802); n=100 |
| Aortic valve | Post: Aortic valve morphology | 0.706 (0.595–0.808); n=97 | 0.595 (0.463–0.717); n=100 | 0.848 (0.753–0.931); n=100 |
| Aortic valve | Post: Aortic regurgitation | 0.840 (0.736–0.925); n=97 | 0.722 (0.594–0.834); n=100 | 0.774 (0.664–0.871); n=100 |
| Aortic valve | Post: Aortic stenosis | 0.908 (0.809–0.978); n=97 | 0.751 (0.617–0.866); n=100 | 0.848 (0.741–0.941); n=100 |
| Aortic valve | Post: Aortic valve type | 0.749 (0.643–0.846); n=97 | 0.836 (0.746–0.921); n=100 | 0.858 (0.767–0.937); n=100 |
| Mitral valve | Pre: Mitral regurgitation | 0.934 (0.870–0.987); n=97 | 0.973 (0.928–1.000); n=100 | 0.972 (0.929–1.000); n=100 |
| Mitral valve | Pre: Mitral stenosis | 0.950 (0.875–1.000); n=97 | 0.894 (0.779–0.977); n=100 | 0.874 (0.757–0.973); n=100 |
| Mitral valve | Pre: Mitral valve type | 0.452 (0.260–0.640); n=97 | 0.465 (0.295–0.627); n=100 | 0.065 (0.000–0.203); n=100 |
| Mitral valve | Post: Mitral regurgitation | 0.853 (0.765–0.932); n=97 | 0.861 (0.777–0.932); n=100 | 0.917 (0.848–0.972); n=100 |
| Mitral valve | Post: Mitral stenosis | 0.765 (0.623–0.889); n=97 | 0.741 (0.591–0.870); n=100 | 0.772 (0.634–0.893); n=100 |
| Mitral valve | Post: Mitral valve type | 0.710 (0.572–0.835); n=97 | 0.606 (0.457–0.737); n=100 | 0.452 (0.272–0.613); n=100 |
| Tricuspid valve | Pre: Tricuspid regurgitation | 0.893 (0.819–0.959); n=97 | 0.969 (0.920–1.000); n=100 | 0.954 (0.897–1.000); n=100 |
| Tricuspid valve | Pre: Tricuspid valve type | 0.900 (0.805–0.979); n=97 | 0.805 (0.690–0.902); n=100 | 0.032 (0.000–0.083); n=100 |
| Tricuspid valve | Post: Tricuspid regurgitation | 0.815 (0.721–0.896); n=97 | 0.864 (0.770–0.941); n=100 | 0.902 (0.829–0.969); n=100 |
| Tricuspid valve | Post: Tricuspid valve type | 0.773 (0.646–0.884); n=97 | 0.698 (0.571–0.816); n=100 | 0.144 (0.065–0.241); n=100 |
| Pulmonic valve | Pre: Pulmonic regurgitation | 0.902 (0.829–0.959); n=97 | 0.889 (0.809–0.957); n=100 | 0.933 (0.868–0.986); n=100 |
| Pulmonic valve | Post: Pulmonic regurgitation | 0.754 (0.643–0.849); n=97 | 0.731 (0.625–0.827); n=100 | 0.868 (0.785–0.943); n=100 |
| Left ventricle | Pre: LV size | 0.911 (0.828–0.981); n=97 | 0.875 (0.763–0.959); n=100 | 0.902 (0.721–1.000); n=99 |
| Left ventricle | Pre: LV systolic function | 0.899 (0.814–0.967); n=97 | 0.976 (0.916–1.000); n=100 | 0.734 (0.551–0.892); n=100 |
| Left ventricle | Pre: Regional wall-motion abnormality | 0.738 (0.593–0.861); n=97 | 0.930 (0.819–1.000); n=100 | 0.908 (0.792–1.000); n=99 |
| Left ventricle | Post: LV size | 0.702 (0.583–0.811); n=97 | 0.614 (0.487–0.738); n=100 | 0.815 (0.669–0.932); n=99 |
| Left ventricle | Post: LV systolic function | 0.849 (0.752–0.929); n=97 | 0.827 (0.710–0.923); n=100 | 0.727 (0.570–0.866); n=100 |

| Module | Variable | A–B $\kappa$ (95% CI); n | A–C $\kappa$ (95% CI); n | A–D $\kappa$ (95% CI); n |
|---|---|---|---|---|
| Left ventricle | Post: Regional wall-motion abnormality | 0.702 (0.567–0.823); n=97 | 0.634 (0.483–0.776); n=99 | 0.760 (0.594–0.893); n=100 |
| Right ventricle | Pre: RV size | 0.868 (0.777–0.946); n=97 | 0.930 (0.842–1.000); n=100 | 0.933 (0.819–1.000); n=100 |
| Right ventricle | Pre: RV systolic function | 0.862 (0.768–0.944); n=97 | 0.949 (0.864–1.000); n=100 | 0.867 (0.596–1.000); n=100 |
| Right ventricle | Post: RV size | 0.801 (0.696–0.890); n=97 | 0.734 (0.601–0.849); n=100 | 0.857 (0.734–0.951); n=99 |
| Right ventricle | Post: RV systolic function | 0.865 (0.775–0.941); n=97 | 0.801 (0.677–0.904); n=100 | 0.886 (0.758–0.974); n=100 |
| Aorta | Pre: Aortic dissection | 0.732 (0.591–0.860); n=97 | 0.351 (0.160–0.526); n=100 | 0.250 (0.128–0.383); n=100 |
| Aorta | Post: Aortic dissection | 0.479 (0.304–0.650); n=93 | 0.488 (0.307–0.658); n=100 | 0.266 (0.120–0.410); n=100 |
| Pericardium | Pre: Pericardial effusion | 0.740 (0.616–0.853); n=97 | 0.873 (0.746–0.971); n=100 | 0.849 (0.717–0.950); n=100 |
| Pericardium | Post: Pericardial effusion | 0.615 (0.480–0.741); n=97 | 0.651 (0.491–0.789); n=100 | 0.716 (0.532–0.869); n=100 |
| Report-level timepoints | TEE timepoints available | 0.867 (0.597–1.000); n=96 | 1.000 (1.000–1.000); n=100 | 0.688 (0.242–1.000); n=100 |

**Note.** CIs use 5,000 paired-report bootstrap resamples. Reviewer assignment was nonrandom and informed by case complexity and experience; comparisons are descriptive, not reviewer-performance tests.

### Table S5. Sampling-Weighted Estimates of Categorical Cohen's Kappa

| Module | Variable | Paired n | Unweighted $\kappa$ | Weighted $\kappa$ | $\Delta\kappa$ (weighted − unweighted) |
|---|---|---|---|---|---|
| Aortic valve | Pre: Aortic valve morphology | 297 | 0.866 | 0.881 | +0.014 |
| Aortic valve | Pre: Aortic regurgitation | 297 | 0.906 | 0.907 | +0.001 |
| Aortic valve | Pre: Aortic stenosis | 297 | 0.932 | 0.936 | +0.004 |
| Aortic valve | Pre: Aortic valve type | 297 | 0.753 | 0.759 | +0.006 |
| Aortic valve | Post: Aortic valve morphology | 297 | 0.719 | 0.720 | +0.001 |
| Aortic valve | Post: Aortic regurgitation | 297 | 0.779 | 0.762 | -0.018 |
| Aortic valve | Post: Aortic stenosis | 297 | 0.834 | 0.810 | -0.024 |
| Aortic valve | Post: Aortic valve type | 297 | 0.816 | 0.833 | +0.017 |
| Mitral valve | Pre: Mitral regurgitation | 297 | 0.960 | 0.967 | +0.008 |
| Mitral valve | Pre: Mitral stenosis | 297 | 0.907 | 0.894 | -0.013 |
| Mitral valve | Pre: Mitral valve type | 297 | 0.364 | 0.338 | -0.026 |
| Mitral valve | Post: Mitral regurgitation | 297 | 0.877 | 0.885 | +0.008 |
| Mitral valve | Post: Mitral stenosis | 297 | 0.760 | 0.759 | -0.001 |
| Mitral valve | Post: Mitral valve type | 297 | 0.601 | 0.564 | -0.037 |
| Tricuspid valve | Pre: Tricuspid regurgitation | 297 | 0.938 | 0.955 | +0.017 |
| Tricuspid valve | Pre: Tricuspid valve type | 297 | 0.558 | 0.430 | -0.128 |
| Tricuspid valve | Post: Tricuspid regurgitation | 297 | 0.862 | 0.877 | +0.016 |
| Tricuspid valve | Post: Tricuspid valve type | 297 | 0.529 | 0.436 | -0.093 |
| Pulmonic valve | Pre: Pulmonic regurgitation | 297 | 0.909 | 0.908 | -0.001 |
| Pulmonic valve | Post: Pulmonic regurgitation | 297 | 0.786 | 0.795 | +0.008 |
| Left ventricle | Pre: LV size | 296 | 0.900 | 0.890 | -0.010 |
| Left ventricle | Pre: LV systolic function | 297 | 0.894 | 0.887 | -0.007 |
| Left ventricle | Pre: Regional wall-motion abnormality | 296 | 0.841 | 0.890 | +0.048 |
| Left ventricle | Post: LV size | 296 | 0.703 | 0.695 | -0.008 |
| Left ventricle | Post: LV systolic function | 297 | 0.818 | 0.796 | -0.022 |

| Module | Variable | Paired n | Unweighted $\kappa$ | Weighted $\kappa$ | $\Delta\kappa$ (weighted − unweighted) |
|---|---|---|---|---|---|
| Left ventricle | Post: Regional wall-motion abnormality | 296 | 0.699 | 0.688 | -0.011 |
| Right ventricle | Pre: RV size | 297 | 0.906 | 0.923 | +0.018 |
| Right ventricle | Pre: RV systolic function | 297 | 0.899 | 0.921 | +0.023 |
| Right ventricle | Post: RV size | 296 | 0.799 | 0.798 | -0.001 |
| Right ventricle | Post: RV systolic function | 297 | 0.854 | 0.850 | -0.004 |
| Aorta | Pre: Aortic dissection | 297 | 0.454 | 0.368 | -0.086 |
| Aorta | Post: Aortic dissection | 293 | 0.405 | 0.395 | -0.011 |
| Pericardium | Pre: Pericardial effusion | 297 | 0.814 | 0.845 | +0.031 |
| Pericardium | Post: Pericardial effusion | 297 | 0.660 | 0.674 | +0.014 |
| Report-level timepoints | TEE timepoints available | 296 | 0.854 | 0.853 | -0.001 |

**Note.** Weights: random UPenn=4.50, random UW=3.75. Three legacy UPenn edge reports were excluded (total weights=922). Weighted kappa uses weighted category marginals; point estimates have no design-based CIs, and reviewer assignments were nonrandom.

### Table S6. Categorical Reliability by Institution and Report Sampling Stratum

| Module | Variable | UPenn complex κ (95% CI); n | UPenn random κ (95% CI); n | UW complex κ (95% CI); n | UW random κ (95% CI); n |
|---|---|---|---|---|---|
| Aortic valve | Pre: Aortic valve morphology | 0.827 (0.658–0.957); n=47 | 0.880 (0.779–0.961); n=100 | 0.810 (0.663–0.935); n=50 | 0.891 (0.801–0.964); n=100 |
| Aortic valve | Pre: Aortic regurgitation | 0.859 (0.730–0.970); n=47 | 0.882 (0.802–0.947); n=100 | 0.925 (0.826–1.000); n=50 | 0.937 (0.875–0.987); n=100 |
| Aortic valve | Pre: Aortic stenosis | 0.878 (0.739–1.000); n=47 | 0.905 (0.817–0.980); n=100 | 0.918 (0.812–1.000); n=50 | 0.958 (0.903–1.000); n=100 |
| Aortic valve | Pre: Aortic valve type | 0.762 (0.537–0.937); n=47 | 0.767 (0.596–0.904); n=100 | 0.644 (-0.010–1.000); n=50 | 0.660 (0.000–1.000); n=100 |
| Aortic valve | Post: Aortic valve morphology | 0.803 (0.643–0.933); n=47 | 0.680 (0.554–0.796); n=100 | 0.560 (0.390–0.723); n=50 | 0.725 (0.595–0.849); n=100 |
| Aortic valve | Post: Aortic regurgitation | 0.905 (0.782–1.000); n=47 | 0.777 (0.668–0.874); n=100 | 0.762 (0.592–0.902); n=50 | 0.715 (0.586–0.826); n=100 |
| Aortic valve | Post: Aortic stenosis | 0.919 (0.796–1.000); n=47 | 0.783 (0.658–0.888); n=100 | 0.855 (0.615–1.000); n=50 | 0.819 (0.697–0.923); n=100 |
| Aortic valve | Post: Aortic valve type | 0.723 (0.556–0.880); n=47 | 0.770 (0.657–0.871); n=100 | 0.727 (0.580–0.871); n=50 | 0.907 (0.828–0.969); n=100 |
| Mitral valve | Pre: Mitral regurgitation | 0.948 (0.867–1.000); n=47 | 0.962 (0.913–1.000); n=100 | 0.913 (0.804–1.000); n=50 | 0.983 (0.942–1.000); n=100 |
| Mitral valve | Pre: Mitral stenosis | 1.000 (1.000–1.000); n=47 | 0.914 (0.759–1.000); n=100 | 0.886 (0.694–1.000); n=50 | 0.855 (0.743–0.950); n=100 |
| Mitral valve | Pre: Mitral valve type | 0.486 (0.236–0.714); n=47 | 0.376 (0.217–0.537); n=100 | 0.286 (0.000–0.627); n=50 | 0.171 (0.000–0.357); n=100 |
| Mitral valve | Post: Mitral regurgitation | 0.869 (0.750–0.973); n=47 | 0.883 (0.806–0.948); n=100 | 0.828 (0.692–0.943); n=50 | 0.891 (0.807–0.967); n=100 |
| Mitral valve | Post: Mitral stenosis | 0.787 (0.606–0.956); n=47 | 0.752 (0.595–0.887); n=100 | 0.679 (0.381–0.895); n=50 | 0.745 (0.611–0.863); n=100 |
| Mitral valve | Post: Mitral valve type | 0.803 (0.637–0.934); n=47 | 0.583 (0.436–0.715); n=100 | 0.411 (0.116–0.681); n=50 | 0.451 (0.265–0.619); n=100 |
| Tricuspid valve | Pre: Tricuspid regurgitation | 0.893 (0.786–0.973); n=47 | 0.970 (0.925–1.000); n=100 | 0.886 (0.766–0.972); n=50 | 0.951 (0.889–1.000); n=100 |
| Tricuspid valve | Pre: Tricuspid valve type | 0.652 (0.000–1.000); n=47 | 0.016 (0.000–0.053); n=100 | 0.545 (0.000–1.000); n=50 | 0.492 (0.211–0.728); n=100 |
| Tricuspid valve | Post: Tricuspid regurgitation | 0.806 (0.666–0.919); n=47 | 0.887 (0.810–0.957); n=100 | 0.817 (0.685–0.924); n=50 | 0.880 (0.788–0.954); n=100 |
| Tricuspid valve | Post: Tricuspid valve type | 0.909 (0.717–1.000); n=47 | 0.083 (0.015–0.168); n=100 | 0.212 (0.000–0.471); n=50 | 0.479 (0.270–0.664); n=100 |
| Pulmonic valve | Pre: Pulmonic regurgitation | 0.769 (0.605–0.903); n=47 | 0.845 (0.749–0.931); n=100 | 1.000 (1.000–1.000); n=50 | 0.942 (0.880–0.986); n=100 |

| Module | Variable | UPenn complex<br>$\kappa$ (95% CI); n | UPenn random<br>$\kappa$ (95% CI); n | UW complex<br>$\kappa$ (95% CI); n | UW random<br>$\kappa$ (95% CI); n |
|---|---|---|---|---|---|
| Pulmonic valve | Post: Pulmonic regurgitation | 0.709 (0.537–0.854); n=47 | 0.756 (0.651–0.854); n=100 | 0.760 (0.604–0.905); n=50 | 0.801 (0.701–0.888); n=100 |
| Left ventricle | Pre: LV size | 0.911 (0.800–1.000); n=47 | 0.871 (0.761–0.958); n=99 | 0.893 (0.707–1.000); n=50 | 0.910 (0.745–1.000); n=100 |
| Left ventricle | Pre: LV systolic function | 1.000 (1.000–1.000); n=47 | 0.877 (0.771–0.973); n=100 | 0.668 (0.384–0.884); n=50 | 0.889 (0.754–1.000); n=100 |
| Left ventricle | Pre: Regional wall-motion abnormality | 0.832 (0.676–0.965); n=47 | 0.912 (0.806–1.000); n=99 | 0.463 (0.131–0.735); n=50 | 0.925 (0.795–1.000); n=100 |
| Left ventricle | Post: LV size | 0.796 (0.650–0.915); n=47 | 0.636 (0.518–0.749); n=99 | 0.437 (0.192–0.656); n=50 | 0.776 (0.584–0.916); n=100 |
| Left ventricle | Post: LV systolic function | 0.944 (0.854–1.000); n=47 | 0.754 (0.628–0.868); n=100 | 0.693 (0.487–0.872); n=50 | 0.830 (0.673–0.943); n=100 |
| Left ventricle | Post: Regional wall-motion abnormality | 0.712 (0.523–0.870); n=47 | 0.622 (0.475–0.756); n=100 | 0.550 (0.206–0.806); n=50 | 0.785 (0.612–0.920); n=99 |
| Right ventricle | Pre: RV size | 0.803 (0.653–0.917); n=47 | 0.939 (0.864–1.000); n=100 | 0.954 (0.836–1.000); n=50 | 0.909 (0.744–1.000); n=100 |
| Right ventricle | Pre: RV systolic function | 0.832 (0.697–0.943); n=47 | 0.975 (0.911–1.000); n=100 | 0.874 (0.688–1.000); n=50 | 0.788 (0.470–1.000); n=100 |
| Right ventricle | Post: RV size | 0.834 (0.696–0.943); n=47 | 0.817 (0.711–0.912); n=99 | 0.725 (0.532–0.892); n=50 | 0.745 (0.563–0.881); n=100 |
| Right ventricle | Post: RV systolic function | 0.837 (0.708–0.944); n=47 | 0.914 (0.826–0.980); n=100 | 0.860 (0.686–1.000); n=50 | 0.748 (0.593–0.881); n=100 |
| Aorta | Pre: Aortic dissection | 0.480 (0.233–0.720); n=47 | 0.659 (0.426–0.841); n=100 | 0.840 (0.668–0.963); n=50 | 0.112 (0.017–0.213); n=100 |
| Aorta | Post: Aortic dissection | 0.306 (0.011–0.578); n=43 | 0.589 (0.375–0.766); n=100 | 0.561 (0.341–0.754); n=50 | 0.226 (0.109–0.353); n=100 |
| Pericardium | Pre: Pericardial effusion | 0.699 (0.519–0.859); n=47 | 0.870 (0.728–0.972); n=100 | 0.779 (0.590–0.926); n=50 | 0.852 (0.730–0.950); n=100 |
| Pericardium | Post: Pericardial effusion | 0.563 (0.379–0.739); n=47 | 0.675 (0.522–0.816); n=100 | 0.649 (0.418–0.841); n=50 | 0.685 (0.504–0.838); n=100 |
| Report-level timepoints | TEE timepoints available | 0.857 (0.574–1.000); n=47 | 0.886 (0.689–1.000); n=100 | NE; n=49 | 0.000 (CI NE); n=100 |

**Note.** All estimates are unweighted Cohen's $\kappa$ from pre-adjudication anchor-versus-assigned-secondary comparisons, with 95% paired-report percentile bootstrap CIs (5,000 resamples). Complex denotes the originally selected edge-case sample. Each cell reports the number of reports with paired nonmissing values for that variable; the four formal strata contain 47 UPenn complex, 100 UPenn random, 50 UW complex, and 100 UW random reports. Three of the original 50 UPenn complex reports were legacy teaching examples excluded from all formal IRR analyses. NE indicates a coefficient that cannot be estimated, typically because of insufficient category variation. CI NE indicates that $\kappa$ was calculated but a 95% bootstrap interval could not be estimated reliably. Strata were deliberately selected rather than randomized; differences are descriptive, not formal site or case-complexity comparisons.

### Supplemental Methods S1. Code and Data Availability

A public repository for this study is available through the TEECompass GitHub organization (https://github.com/TEECompass/tee-reference-standard-v1). Analysis code is being prepared for release, subject to institutional review of licensing and intellectual property. Patient-level clinical reports and adjudication data are not publicly available due to institutional data-use agreements and privacy protections.
